# From peas to people - using quantitative traits to aid genetic discovery in depression

**DOI:** 10.1101/2024.09.12.24313543

**Authors:** Lynsey S. Hall, Mark J. Adams, Yanni Zeng, Jude Gibson, Ella M. Wigmore, Ana Maria Fernandez-Pujals, Generation Scotland, Heather C. Whalley, Chris S. Haley, Andrew M. McIntosh

## Abstract

A key component of Mendel’s work is what we now refer to as pleiotropy - when variation in one gene gives rise to variation in multiple phenotypes.

This study focuses on aiding genetic discovery in depression by revisiting the depressed phenotype and developing a quantitative trait in a large mixed family and population study, using analyses built upon the theory which underpins Mendel’s pleiotropic observations - the relationship between phenotypic variation and genetic variation.

Measures of genetic covariation were used to evaluate and rank ten measures of mood, personality, and cognitive ability as endophenotypes for depression. The highest-ranking traits were subjected to principal component analysis, and the first principal component used to create multivariate measures of depression.

Four traits fulfilled most endophenotype criteria, however, only two traits (neuroticism and the general health questionnaire) consistently ranked highest across all measures of covariation. As such, three composite traits were derived incorporating two, three, or four traits.

Composite traits were compared to the binary classification of depression and to their constituent univariate traits in terms of their coheritability, their ability to identify risk loci in a genome-wide association analysis, and phenotypic variance explained by polygenic profile scores for depression.

Association analyses of binary depression, univariate traits, and composite traits yielded no genome-wide significant results. However, composite traits were more heritable and more highly correlated with depression than their constituent traits, suggesting that analysing candidate endophenotypes in combination captures more of the heritable component of depression and may in part be limited by sample size in the current study.

## Introduction

If I were to ask you, “How do ornamental flowers relate to depression?”, you’d be forgiven for thinking that perhaps a floral gift in a bid to bring cheer was what I had in mind. Whilst flowers will always be graciously received by Hall and colleagues, that is not what I am referring to here. I am, in fact, referring to celebrated geneticist Gregor Mendel and his experiments in plant hybridization. The link being the concept of pleiotropy.

The term pleiotropy (or “pleiotropie”, derived from the Greek meaning “more ways”) was coined in 1910 by German geneticist Ludwig Plate, who defined it as a phenomenon in which a single locus affects two or more distinct phenotypic traits. Forty-four years prior to this neologism, Mendel described what would later go on to be called pleiotropy. In experimenting with the artificial fertilization of ornamental pea plants to obtain new colour variations, he noted that three of the seven characteristics measured (seed coat colour, flower petal colour, and axial spots) consistently co-segregated. Mendel therefore considered these traits correlated and under the control of a single “factor” or gene as we would say today (Mendel, 1866; Stearns, 2010).

Perhaps it seems an unusual choice, linking a paper which explores the genetic basis of depression to the founder of Mendelian genetics. For if there is one thing depression is not - it is Mendelian. However, neither were the traits observed in Mendel’s pleiotropic peas! This is because Mendel’s experiments dealt with binary traits. The peas were yellow or green, wrinkled or round. This dichotomization obscured a more nuanced phenotypic truth, which was uncovered in a repeat of the hybridization experiments by biometrician Raphael Weldon. Weldon noted that the peas were not unambiguously yellow and green but sat on a continuum of colour from pure yellow, through various intermediate yellow-green shades, to pure green (Weldon, 1902).

Weldon drew three primary conclusions from this experiment, namely that variation, ancestry, and environment are important. We lose valuable information about the genetic architecture of a trait if we categorize a quantitative phenotype as a binary, if we focus only on parent-offspring phenotypic resemblance without considering more detailed pedigrees and deeper ancestries, and if we ignore the environmental context (both exogenous and endogenous) within which a genetic factor resides and interacts (Radick, 2016).

Moving from peas to people - in studies of disease, people are categorized in a binary manner as being either with or without illness. Whilst this is effective for Mendelian traits with dominant inheritance patterns, for polygenic traits like depression we run into the same classification problems as were uncovered by Weldon. The phenotypic reality is that people are not simply ill or well, but on a spectrum of illness, with the genetic liability for disease most likely being quantitative in distribution also (Yang et al., 2010; Tenesa and Haley, 2013). By classifying a quantitative liability as a binary trait we lose substantial information on trait variation, resulting in a reduction in statistical power to detect genetic effects. This is particularly problematic in a trait like depression where genetic discovery is already limited by the trait’s modest heritability (Sullivan et al., 2000; Fernandez-Pujals et al., 2015; Howard et al., 2019), as heritability is the upper bound of what we can hope to interrogate as geneticists (Tenesa and Haley, 2013).

The purpose of the current study is to use information from measures of mood, personality, and cognitive ability which are genetically correlated with a diagnosis of depression to derive a quantitative depression phenotype that maximizes the variance and heritability, with a view to improving the statistical power to identify genetic risk variants.

For this, we used data from the Generation Scotland cohort (Smith et al., 2006; Smith, Campbell, et al., 2013). This sample is well-suited for such an analysis as it has detailed phenotype information on ten well-validated quantitative measures and a structured clinical diagnosis of depression in the same individuals, in addition to being a substantial sample size (N∼24,000) including a mixture of different familial relationships and unrelated population members.

We selected quantitative traits to take forward for multivariate analysis by using the endophenotype criteria outlined by Gottesman and Gould. These criteria state that traits should be heritable, genetically and phenotypically correlated with the trait of interest, state independent, co-segregating with illness in families, and observed at a higher rate in unaffected relatives than in unrelated controls (Gottesman and Gould, 2003).

Univariate and multivariate traits were evaluated using the endophenotype ranking value (or absolute coheritability) (Glahn et al., 2012), and phenotypic variance explained by polygenic risk scores for depression. Given the latter method uses only common trait-associated variants, selecting quantitative traits for further genetic analysis based on this method may prove advantageous for gene-discovery in genome-wide association analysis.

Whether our quantitative measures of depression had improved statistical power relative to the binary case/control classification was assessed by performing a genome-wide association analysis of each trait and establishing whether quantitative traits had an increased number of extreme test statistics (P≤1E-05) or identified any genomic regions of interest beyond those seen in binary depression.

## Materials and Methods

### Sample description

Analyses were conducted using data from the Generation Scotland: The Scottish Family Health Study cohort (GS), and UK Biobank (UKB) under UKB data application number 4844. GS received ethical approval from the NHS Tayside Committee on Medical Research Ethics (REC Reference Numbers: 05/S1401/89 and 10/S1402/20). UKB received ethical approval from the Research Ethics Committee (REC Reference Number: 11/NW/0382).

### Generation Scotland: The Scottish Family Health Study

GS is a family- and population-based study consisting of 23,690 adult participants recruited via general medical practices across Scotland. Participants were not ascertained on the basis of having any particular disorder. The recruitment protocol and sample characteristics are described in detail elsewhere(Smith et al., 2006; Smith, Campbell, et al., 2013).

### Depression phenotype

A diagnosis of depression (MDD) was made using the structured clinical interview for DSM-IV disorders (SCID), (First et al., 1997). After exclusion criteria and phenotypic refinement, as described in Hall et al (Hall et al., 2018) there were 2,603 depression cases and 16,122 controls.

### Candidate endophenotypes

Ten measures of mood, personality, and cognition were tested for adherence to endophenotype characteristics to take forward for multivariate analysis.

Current levels of MDD and psychological distress were assessed with the 28-item General Health Questionnaire (GHQ), using the Likert scoring method (Goldberg and Hillier, 1979). Participants were screened for symptoms of bipolar spectrum disorders using the Mood Disorder Questionnaire (MDQ) (Hirschfeld et al., 2000; Hirschfeld, 2002), and schizotypal features using the schizotypal personality questionnaire-brief (SPQ), (Raine and Benishay, 1995). The personality traits of neuroticism (EPQN) and extraversion (EPQE) were assessed using the relevant sections of the Eysenck Personality Questionnaire (Eysenck et al., 1985; Eysenck and Eysenck, 1994). Verbal declarative memory was assessed using immediate (LM1) and delayed (LM2D) recall with the Wechsler logical memory test (Wechsler, 1997a). Executive function was assessed using the letter-based phonemic verbal fluency (VF) test (Raven et al., 1998). Processing speed was measured using the Wechsler digit symbol substitution task (DSC) (Wechsler, 1997b). Acquired verbal knowledge and vocabulary were assessed using the Mill Hill Vocabulary Scale (MHV) (Raven et al., 1998). Summary statistics of candidate endophenotypes are shown in Table 1.

**Table 1.** Summary statistics for mood, personality, and cognitive traits. N: number of individuals with data for each phenotype; Max. Score: maximum theoretical score for each phenotype; Range: range of scores obtained for each phenotype; Mean (sd): mean and standard deviation for each phenotype.

| Phenotype | N | Max. Score | Range | Mean (sd) |
| --- | --- | --- | --- | --- |
| DSC | 19,765 | 133 | 0-133 | 72.2 (17.2) |
| EPQE | 19,866 | 12 | 0-12 | 7.8 (3.5) |
| EPQN | 19,874 | 12 | 0-12 | 3.9 (3.2) |
| GHQ | 19,693 | 84 | 0-84 | 16.0 (8.9) |
| LM1 | 19,855 | 25 | 0-25 | 16.1 (3.9) |
| LM2D | 19,751 | 25 | 0-25 | 14.9 (4.3) |
| MDQ | 10,357 | 13 | 0-13 | 2.5 (3.2) |
| MHV | 19,646 | 44 | 0-44 | 30.1 (4.7) |
| SPQ | 10,845 | 22 | 0-22 | 3.9 (3.7) |
| VF | 19,760 | - | 0-97 | 39.7 (11.7) |

Impossible values, as dictated by each trait’s maximum theoretical score (Smith, Campbell, et al., 2013), were set to missing. Phenotypes were transformed to improve normality using the Box-Cox power transformation for linear models (Box and Cox, 1964), implemented in the MASS package (Venables and Ripley. B. D., 2002) in R (R Development Core Team, 2016). To simplify the interpretation of effect sizes the transformed traits were standardized to have a mean of zero and a unit standard deviation using Y=(X-μ)/σ.

### Genetic data

Autosomal genotype data were available for 19,994 individuals in GS. Quality control (QC) procedures are described in detail in Hall et al (Hall et al., 2018). In brief, QC exclusion thresholds of □≥□3% individual missingness, SNP call rate of ≤98%, Hardy Weinberg Equilibrium (HWE) P-value of □≤1E-06, and minor allele frequency (MAF) of ≤□1% were employed. At the time of analysis, imputed genetic data was not available, resulting in 561,125 SNPs for use in analysis.

### UKB

UKB is a health research resource of ∼500,000 middle-aged participants (aged 40 to 73 years) recruited from the UK that aims to improve the prevention, diagnosis, and treatment of a wide range of illnesses. The recruitment protocol and sample characteristics are described in detail elsewhere (Sudlow et al., 2015).

### Depression phenotype

Current and previous depressive symptoms were assessed by items relating to the lifetime experience of minor and major depression, items from the Patient Health Questionnaire and items on help-seeking for mental health. Case status was defined as either ‘probable single lifetime episode of major depression’ or ‘probable recurrent major depression (moderate and severe)’. Individuals with probable bipolar disorder or mild depressive/manic symptoms were excluded. In the current study, we employ a “strict” definition of MDD where cases (N=35,960) were defined as single, moderate, or recurrent MDD, and controls (N=91,879) were defined as an absence of putative MDD. A detailed description of the UKB putative depression phenotype is given in Smith et al (Smith, Nicholl, et al., 2013).

### Genetic data

Autosomal genotype data was available for all individuals with our MDD phenotype, all of whom had European ancestry. We excluded participants who were also in GS, their relatives, and relatives of remaining UKB participants (≤3rd degree), were identified by a kinship coefficient ≥0.0442 using the KING toolset (Manichaikul et al., 2010). The genotyping and imputation process is described in detail elsewhere (UK Biobank, 2015). The analyses presented here were restricted to autosomal variants with an imputation INFO score□≥□0.9, MAF□≥□0.5%, and array genotype missingness < 0.02.

### Statistical analyses

#### Genome-wide Association Analysis (GWAA)

GWAA of MDD, univariate and multivariate endophenotypes using GS data were conducted using mixed linear model-based association (MLMA) analysis (Yang et al., 2014) implemented in GCTA (v1.25.) (Yang et al., 2011). Age, age^2^, and sex were fitted as fixed effects. Two genomic relationship matrices (GRMs), created using the mixed linear model with candidate marker excluded approach (Yang et al., 2014), were fitted as random effects to account for the enrichment of familial relationships in GS (Zaitlen et al., 2013). The first GRM included pairwise relationship coefficients for all individuals. The second GRM had off-diagonal elements□<□0.05 set to 0.

As MLMA operates on a linear scale, for the binary MDD analysis betas and their corresponding standard errors were transformed to odds ratios and 95% confidence intervals on the liability scale using a Taylor transformation expansion series (Cortes et al., 2013; Visscher et al., 2014).

GWAA of MDD using UKB data was conducted using logistic regression, implemented in PLINK v1.9 (Chang et al., 2015). Age, age^2^, sex, assessment centre, genotyping array, batch, and the first 15 PCs fitted as fixed effects.

#### Polygenic Risk Score Analysis (PRS)

Polygenic risk scores for MDD were created PLINK, in accordance with methods described by Purcell et al (Purcell et al., 2009). MDD GWAA summary statistics from the Psychiatric Genomics Consortium (Sullivan et al., 2013) were used to provide marker weights and evidence of association for SNPs. PRS were generated at five p-value thresholds (P_T_□<□0.01,□<□0.05,□<□0.1,□<□0.5 and all SNPs). SNPs common to PGC and GS datasets were extracted from GS and subjected to clump-based linkage disequilibrium (LD) pruning, using an LD R^2^ cut off of 0.25, 300kb sliding window and variance inflation factor of 2. PRS were then standardized using Y=(X- μ)/σ. Variance in trait value explained by polygenic profile score (R^2^) was estimated by multiplying the profile score by its corresponding regression coefficient and estimating its variance. This value was then divided by the variance of the observed phenotype to yield a coefficient of determination between 0 and 1 (Nakagawa and Schielzeth, 2013), and multiplied by 100 to convert R^2^ to percentage variance explained.

#### Testing endophenotype criteria in GS

Only phenotype data from genotyped individuals (n=19,994) was used in these analyses. Where required, to reduce confounding caused by familial relationships, individuals were grouped by affection status into cases (n=2,659), unaffected first-degree relatives (n=2,106), and unrelated controls (n=11,898).

To ensure that common environment was adequately modelled (given the mixed population and family study design of GS), models incorporating shared parent-offspring, sibling, and spousal environmental components as additional random effects were tested using a stepwise likelihood ratio test (LRT) approach. Further details of how environmental components were constructed are provided in the Supplementary Methods of Hall et al (Hall et al., 2018).

All mixed linear model analyses conducted in ASReml-R fitted age, age^2^, and sex as fixed effects, and an inverted additive relationship matrix using pedigree kinship information, common household, and sibling environment as random effects. Any additional model specifications are outlined in the relevant analyses’ descriptions.

#### Associated with illness in the population

The effect of affection status on mean trait scores was calculated using the Wald Conditional F-test (Kenward and Roger, 1997), implemented in ASReml-R (Butler et al., 2007). Standardized traits were fitted as the dependent variable, and affection status was fitted as a fixed effect factor with unrelated controls as the reference group. A conservative Bonferroni corrected P-value threshold of 3.85E-03 was used to indicate statistical significance. Traits were deemed significantly associated with MDD in the population if the mean trait difference between cases and unrelated controls survived multiple testing correction.

#### More extreme in unaffected relatives than in the general population

This endophenotype criteria is an extension of that described in the section above, with the additional requirement that differences in trait mean between unaffected first-degree relatives and unrelated controls also had to survive multiple testing correction and be in the same direction as cases.

#### State independence

It was not possible to explicitly test state independence, as repeated measures were not available in GS at the time of analysis, and the majority of cases (∼80%) were currently depressed at the time of assessment. This introduces collinearity between case/control status and current depression. Therefore, whether the endophenotype was more extreme in unaffected relatives than the general population was used as a proxy measure of state independence (Glahn and Blangero, 2011).

#### Heritability of endophenotypes

Heritability estimates for candidate endophenotypes were estimated using pedigree data and genomic data. Pedigree-based estimates were derived used restricted maximum likelihood methods, implemented in ASReml-R (Butler et al., 2007). As this analysis used theoretical relationships, principal components were not fitted to adjust for population stratification.

#### Genetic and phenotypic correlation

Bivariate analyses were conducted in ASReml-R. Genetic correlations (r_g_) between traits and MDD were calculated using r_g_=cov_A_/√(V_Ai_*V_Aj_) where cov_A_ is the additive covariance, V_Ai_ is the additive variance of the quantitative trait and V_Aj_ is the additive variance of MDD. The significance of r_g_ was calculated by generating a null model assuming no covariance between traits, comparing the alternative and null models using an LRT, the output of which was assessed against a mixed 0.5(χ^2^)+0.5(0) distribution (Visscher, 2006). Similarly, phenotypic correlations (r_p_) between traits and MDD were calculated using r_p_=cov_P_/√(V_Pi_*V_Pj_). However, as ASReml-R does not provide a test statistic for phenotypic correlations, the significance of r_p_ was estimated from the ratio of the phenotypic covariance component to its standard error compared to zero using a Z-test.

### Multivariate analysis

Three multivariate endophenotypes were derived incorporating the two, three, or four traits which ranked most highly across endophenotype criteria. The two-trait measure (“SoS” - sum of Z-scores) was constructed by summing the two standardized variables, as the contributing trait weights cannot be estimated via PCA for fewer than three traits, so this method assigns equal weight to each trait.

Composite traits incorporating three (Gen3) or four (Gen4) endophenotypes were generated by performing spectral decomposition of the genetic correlation matrix (using ASReml r_g_ estimates) using the eigen function in R to compute eigenvalues and eigenvectors. The loading for the first principal component of each trait was then extracted, multiplied by its corresponding standardized trait value, and the resulting values summed together across traits to create the composite trait. This is visualized in Figure 1.

**Figure 1.**
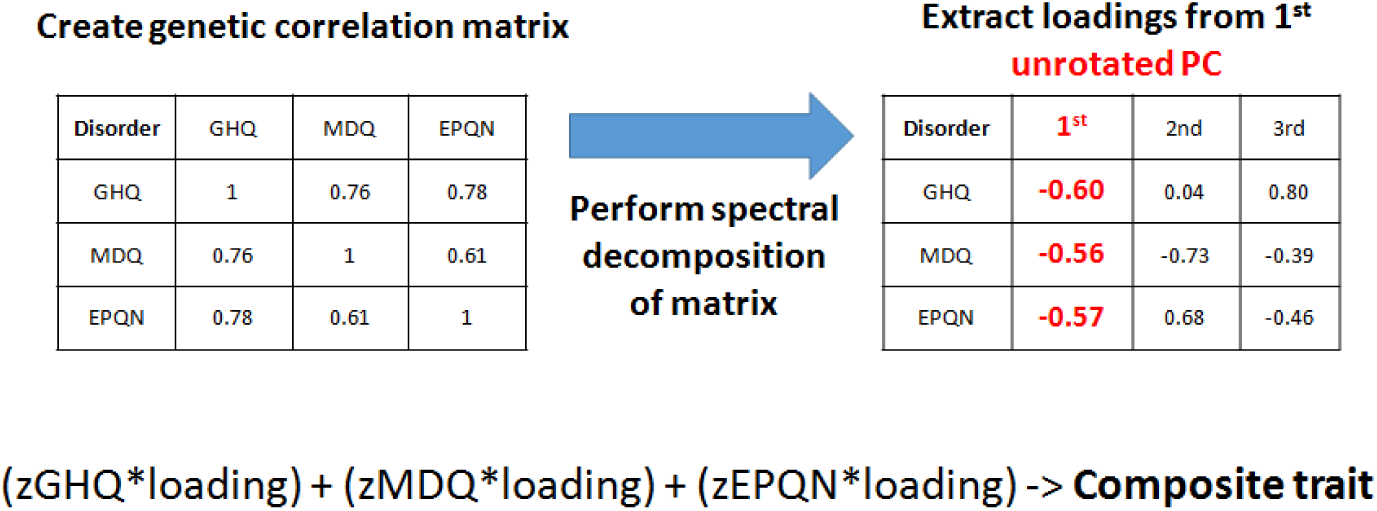
The process by which composite traits using genetic correlations were derived, using Gen3 as an example.

Composite traits were then subjected to the same set of analyses as univariate traits to assess their performance as endophenotypes, relative to their constituent univariate traits.

### Testing performance of endophenotypes

#### Using coheritability

Coheritability (h_ij_) between traits and MDD was calculated using the Endophenotype Ranking Value (Glahn et al., 2012), defined as: h_ij_= |√h ^2^√h ^2^r | where h ^2^ is the estimated heritability of the quantitative trait, h_j_ ^2^ is the estimated heritability for MDD, and r_g_ is the genetic correlation between trait and MDD.

#### Using variance explained by MDD PRS

Traits which had a significant association with MDD PRS surviving a conservative Bonferroni correction (P<7.14E-04) were proposed to be candidate endophenotypes for MDD. Variance explained by PRS was used to rank traits.

#### Using GWAA

The number of test statistics with more extreme associated p-values (P<1E-05) in GWAA of each trait will be used as a metric of whether quantitative candidate endophenotypes have improved power to identify genetic risk variants relative to the binary classification of MDD.

To assess whether quantitative traits provided additional biological insight relative to binary MDD, SNPs with P≤1E-05 which overlapped with a known gene were further explored using the GeneCards database (www.genecards.org) to assess the gene(s)’ function and known disease associations. GeneCards draws its gene-disease associations from multiple external sources, including: OMIM, ClinVar, Orphanet, UniProtKB/Swiss-Prot, Genetic Testing Registry, miR2Disease, LncRNADisease, the University of Copenhagen DISEASES database and Novoseek.

### Replication in UKBiobank

To assess whether univariate and multivariate endophenotypes had a genetic correlation with MDD status in an independent sample, GWAA summary statistics of endophenotypes were assessed against GWAA summary statistics of MDD in UKB using the “rg” function LD Score regression (Bulik-Sullivan, Finucane, et al., 2015b). Prior to bivariate analysis, GWAA summary statistics were reformatted using the “munge” function (Bulik-Sullivan et al., 2015) with pre-computed LD scores estimated from the European-ancestry samples in the 1000 Genomes Project (Altshuler et al., 2012).

## Results

### Selecting endophenotypes for use in multivariate analysis Effect of affection status on trait mean

Six out of ten traits demonstrated a significant effect of case status on mean trait score. Four traits demonstrated an increased trait value in cases: EPQN (Z=36.74, p=3.85e-285), GHQ (Z=34.85, p=2.47e-255), MDQ (Z=19.08, p=2.33e-78), and SPQ (18.47, p=7.69e-74). Two traits demonstrated a decreased trait value in cases: EPQE (Z=-10.32, p=1.01e-24), and DSC (Z=-10.28, p=1.38e-23). Of these six traits, five traits (EPQN, GHQ, SPQ, DSC and MDQ) were also more extreme in both unaffected relatives and cases, although arguably, MDQ showed only nominal significance in unaffected relatives. The direction of effect size was consistent between unaffected relatives and case status. More detailed summary statistics are shown in Supplementary Table 1.

### Heritability of endophenotypes

All traits were moderately heritable, with heritability estimates (h^2^) ranging from h^2^(se) = 0.43(0.03) for DSC to h^2^(se) = 0.11(0.03) for GHQ. Traits with the highest heritability were measures of cognitive ability, followed by personality traits and measures of mood. The observed heritability of MDD was estimated as h^2^(se)=0.17(0.02) which, as would be expected, is markedly lower than the estimate on the liability scale of h^2^(se) = 0.44(0.04) (Fernandez-Pujals et al., 2015). Heritability estimates using pedigree-data are shown in Supplementary Table 2.

### Genetic and phenotypic correlation with MDD

Four traits had a strong (r_g_>0.5) and highly significant (p<1E-10) genetic correlation with MDD - MDQ, GHQ, SPQ, and EPQN. Whilst all of these traits also had a moderate (r_p_>0.19) phenotypic correlation with MDD, this was only statistically significant for two of the traits - GHQ and EPQN. Two traits, DSC and EPQE, had a moderate genetic correlation with MDD, although EPQE did not survive multiple testing correction and both traits had low, non-significant phenotypic correlations.

The remaining four traits, LM1, LM2D, MHV and VF, had low, non-significant genetic and phenotypic correlations with MDD. Results are shown in Supplementary Table 3.

Two traits, GHQ and EPQN, fulfilled all endophenotype criteria. A further three traits, DSC, MDQ and SPQ, fulfilled all endophenotype criteria except sharing a statistically significant phenotypic correlation with MDD. A summary of how well selected quantitative traits fulfilled endophenotype criteria is shown in Table 2.

**Table 2.** Summary of how well selected quantitative traits fulfilled endophenotype criteria. Assoc with illness in pop: the trait is associated with MDD in the population; More extreme in 1st deg rel: the trait is more extreme in first degree relatives; State indep: the trait is state independent; Heritable: the trait is heritable; r_g_: the trait is genetically correlated with MDD; r_p_: the trait is phenotypically correlated with MDD; Y/N: Yes/No. Traits which fulfil most or all of these criteria are highlighted in bold. Composite traits are highlighted in italics.

| Trait | Assoc with illness in pop | More extreme in 1st deg rel | State indep | Heritable | $r_g$ | $r_p$ |
| --- | --- | --- | --- | --- | --- | --- |
| DSC | Y | Y | Y | Y | Y | N |
| EPQE | Y | N | N | Y | N | N |
| **EPQN** | **Y** | **Y** | **Y** | **Y** | **Y** | **Y** |
| **GHQ** | **Y** | **Y** | **Y** | **Y** | **Y** | **Y** |
| LM1 | N | N | N | Y | N | N |
| LM2D | N | N | N | Y | N | N |
| **MDQ** | **Y** | **Y** | **Y** | **Y** | **Y** | **N** |
| MHV | N | N | N | Y | N | N |
| **SPQ** | **Y** | **Y** | **Y** | **Y** | **Y** | **N** |
| VF | N | N | N | Y | N | N |
| *SoS* | Y | Y | Y | Y | Y | Y |
| *Gen3* | Y | Y | Y | Y | Y | Y |
| *Gen4* | Y | Y | Y | Y | Y | Y |

Although DSC is a possible candidate and generally survived multiple testing correction, there was a notable drop in strength of association between DSC and MDD relative to other high-ranking traits. Therefore, only EPQN, GHQ, MDQ and SPQ were taken forward for multivariate analysis.

### Multivariate endophenotypes

Three composite traits were derived, incorporating information from two (“SoS”), three (“Gen3”) and four (“Gen4”) candidate endophenotypes. Composite traits were assessed to ensure they also fulfilled endophenotype criteria, and to compare their criteria values against those from their constituent univariate traits. Univariate and composite traits were then ranked using coheritability and compared to binary MDD in terms of variance explained by MDD PRS and the number of SNPs with a GWAA p-value of <1E-05.

### Effect of affection status on trait mean

All composite traits were significantly more extreme in both unaffected relatives and cases, with the direction of effect size remaining consistent between unaffected relatives and case status. Two traits demonstrated an increased trait value in cases: SoS (Z=41.72, p<3.85e-285) and Gen3 (Z=31.30, p=2.90e-203), whilst Gen4 demonstrated a decreased trait value in cases: Gen4 (Z=-30.16, p=2.77e-190).

Relative to single quantitative traits, only SoS performed better than all of its constituent traits (GHQ and EPQN). Gen3 and Gen4 demonstrated more extreme absolute values than MDQ and SPQ, but not GHQ and EPQN. More detailed summary statistics are shown in Supplementary Table 1.

### Heritability of endophenotypes

All composite traits were moderately heritable. Both Gen3 and Gen4 had a heritability estimate of h^2^ (se) = 0.30 (0.05), higher than the heritability estimates for any of their constituent single traits: EPQN = 0.24 (0.03), MDQ = 0.21 (0.05), SPQ = 0.19 (0.04), and GHQ = 0.11 (0.03). The heritability of SoS = 0.23 (0.03) sat between its constituent traits, albeit much closer to the heritability of EPQN than GHQ. Heritability estimates are shown in Supplementary Table 2.

### Genetic and phenotypic correlation with MDD

All composite traits had high, statistically significant genetic correlations (r_g_) with MDD, and moderate phenotypic correlations which all survived multiple testing correction. The genetic correlation of SoS with MDD, r_g_ (se) = 0.73 (0.11), sat between the values observed for its constituent traits, EPQN (r_g_ = 0.54) and GHQ (r_g_ = 0.84).

However, the phenotypic correlation value, r_p_ (se) = 0.30 (0.007), exceeded constituent traits EPQN (r_p_ = 0.26) and GHQ (r_p_ = 0.25). The genetic correlation estimates for Gen3, r_g_ (se) = 0.90 (0.11) and Gen4, r_g_ (se) = -0.90 (0.11) exceeded all their constituent traits except MDQ, r_g_=0.92 (0.18). The phenotypic correlation estimates for Gen3, r_p_ (se) = 0.32 (0.009) and Gen4, r_p_ (se) = -0.31 (0.009) were more extreme than single quantitative traits. Results are shown in Supplementary Table 3.

In summary, all composite traits fulfilled all endophenotype criteria, in some instances to a more extreme extent than their constituent traits. Interestingly, given the traits were generated using information from matrices of genetic correlation values, phenotypic correlation estimates with MDD were particularly affected. This is also interesting as two of the composite traits (Gen3 and Gen4) included univariate traits (MDQ and SPQ) which did not demonstrate a significant phenotypic correlation with MDD themselves.

### Ranking candidate endophenotypes

Endophenotypes were ranked using three methods (coheritability, variance explained by MDD PRS, and biological insights from GWAA) to try and achieve a holistic overview as to whether genetically correlated univariate and composite traits could yield biological insight beyond that of a binary classification of MDD. Although the focus of this paper is on the multivariate traits and the traits used to create them, all ten univariate traits and three composite traits are considered in the ranking process and presented here for completeness.

### Using coheritability

There was a very strong correlation between coheritability estimates and genetic correlation with MDD (r = 0.94, p = 2.41e-06). Results for coheritability estimates are presented in Table 3. All composite traits and five univariate traits (MDQ, SPQ, EPQN, GHQ, and DSC) remained significant after multiple testing correction. No traits had a higher coheritability than MDD on the liability scale (h_ij_ = 0.44). Using the observed heritability estimate of MDD in the coheritability calculation (h_ijo_) did not change the ranking of traits, but the h_ijo_ estimates are much closer in value to the observed heritability estimate for MDD, h^2^ (se) = 0.17 (0.02). Both Gen3 and Gen4 have a coheritability on the observed scale which is higher than the observed heritability estimate of MDD. This suggests that Gen3 and Gen4 are better predictors of disease liability than the binary definition of MDD. Furthermore, all composite traits have higher coheritability estimates of the liability and observed scale relative to their constituent traits.

**Table 3.** Coheritability estimates for depression, candidate univariate and multivariate endophenotypes on the liability scale (h_ij_) and observed scale (h_ijo_) with their associated P-value, ordered by liability scale value.

| Phenotype | $h_{ij}$ | $h_{ij\circ}$ | P-value |
| --- | --- | --- | --- |
| MDD | 0.44 | 0.17 | - |
| Gen3 | 0.33 | 0.20 | <1E-10 |
| Gen4 | 0.33 | 0.20 | <1E-10 |
| MDQ | 0.25 | 0.15 | <1E-10 |
| SoS | 0.23 | 0.14 | <1E-10 |
| SPQ | 0.19 | 0.12 | <1E-10 |
| EPQN | 0.18 | 0.11 | <1E-10 |
| GHQ | 0.15 | 0.09 | <1E-10 |
| DSC | 0.10 | 0.06 | 5.93E-05 |
| EPQE | 0.06 | 0.03 | 0.008 |
| LM1 | 0.03 | 0.02 | 0.1 |
| VF | 0.02 | 0.01 | 0.26 |
| LM2D | 0.02 | 0.01 | 0.25 |
| MHV | 0.02 | 0.01 | 0.24 |

### Using variance explained by MDD PRS

Analysis was conducted at using SNPs from five different discovery GWAA P-value thresholds (P_T_). As P_T_≤0.5 explained the most variance in the discovery PGC MDD GWAA (Sullivan et al., 2013), this P_T_ will be summarized here (Figure 2, Table 4). Results for all P_T_ are shown in Supplementary Table 4.

**Figure 2.**
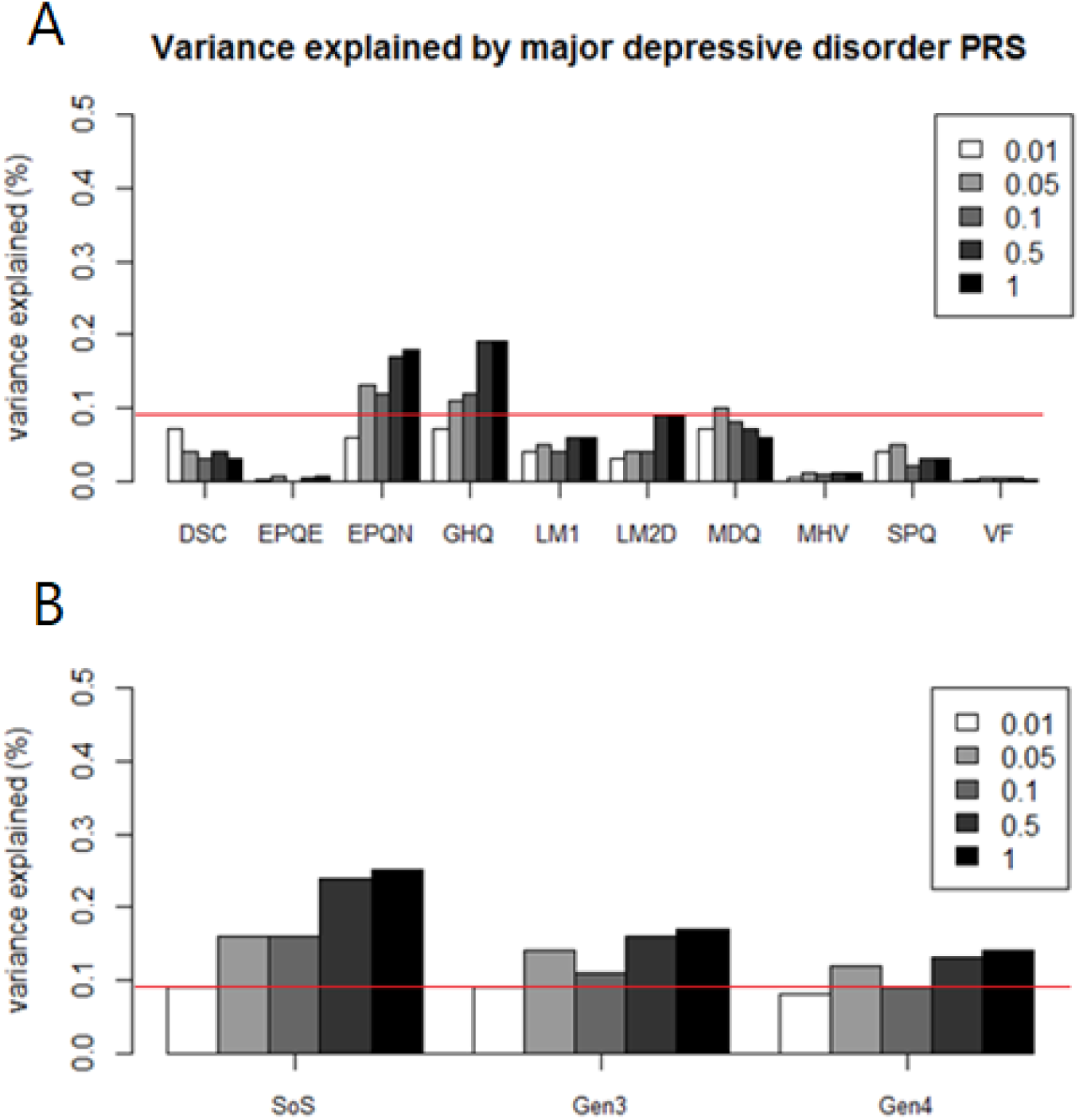
Percentage trait variance explained by MDD PRS at five p-value thresholds (P_T_≤1, P_T_ ≤0.5, P_T_ ≤0.1, P_T_ ≤0.05, P_T_ ≤0.01) in (A) candidate endophenotypes and (B) composite traits. The horizontal line denotes the maximum variance explained by MDD PRS in the binary MDD phenotype.

**Table 4.** Traits ranked by variance explained by depression polygenic risk score at discovery GWAS P_T_≤0.5. P-values which survived multiple testing correction are highlighted in bold.

| Trait | $z\beta/OR$<br>(SE/CI) | Z-Score | % variance<br>explained | P-value |
| --- | --- | --- | --- | --- |
| **SoS** | **0.084**<br>(0.013) | **6.49** | **0.24** | **9.00E-11** |
| **GHQ** | **0.043**<br>(0.008) | **5.74** | **0.19** | **9.85E-09** |
| **EPQN** | **0.041**<br>(0.007) | **5.53** | **0.17** | **3.22E-08** |
| **Gen3** | **0.053**<br>(0.014) | **3.90** | **0.16** | **9.79E-05** |
| Gen4 | -0.053<br>(0.015) | -3.45 | 0.13 | 5.66E-04 |
| LM2D | -0.029<br>(0.007) | -3.98 | 0.09 | 7.03E-05 |
| MDD | 1.09<br>(1.04,1.14) | 3.84 | 0.08 | 1.26E-04 |
| MDQ | 0.026 (0.010) | 2.56 | 0.07 | 0.01 |
| LM1 | -0.024<br>(0.007) | -3.17 | 0.06 | 0.002 |
| DSC | -0.019<br>(0.006) | -2.89 | 0.04 | 0.004 |
| SPQ | 0.017 (0.010) | 1.70 | 0.03 | 0.09 |
| MHV | -0.011<br>(0.007) | -1.60 | 0.01 | 0.11 |
| EPQE | -0.006<br>(0.008) | -0.84 | 0.004 | 0.40 |
| VF | -0.005<br>(0.008) | -0.66 | 0.003 | 0.51 |

PRS for MDD were positively associated with MDD itself, EPQN, GHQ, MDQ, SPQ, SoS and Gen3, and negatively associated with DSC, EPQE, LM1, LM2D, MHV, VF and Gen4. Five traits (three univariate and two composite) survived multiple testing correction (P≤7.14E-04): GHQ, EPQN, LM2D, SoS, and Gen3. Variance explained by MDD PRS for these traits (range 0.09% - 0.24%) was smaller than the variance explained in the PGC MDD mega-analysis (0.6%) (Sullivan et al., 2013), but still (with the exception of LM2D) 2-3 times greater than the variance explained in GS binary MDD (0.08%). Of the composite traits, only SoS (0.24%) outperformed its constituent traits (GHQ = 0.19%, EPQN = 0.17%). This suggests that some quantitative traits are better capturing the polygenic component of MDD than the case/control classification as evidenced by association between score and trait, but predictive capacity is negligible.

### Genome-wide association analysis

GWAA was performed to assess whether univariate or composite quantitative traits associated with MDD have improved power to identify genetic risk variants relative to the binary classification of MDD.

No GWAA analysis yielded any genome-wide significant associations. Forty-four SNPs had a GWAA P-value ≤ 1E-05. The binary classification of MDD had 6 SNPs with a P-value ≤ 1E-05. Two quantitative traits had a higher number of SNPs with more extreme p-values, GHQ (8 SNPs) and SPQ (8 SNPs). Two quantitative traits also had 6 SNPs, MDQ and Gen3. The remaining quantitative traits had fewer SNPs than binary MDD, Gen4 (5 SNPs), EPQN (3 SNPs), and SoS (2 SNPs). This suggests that quantitative traits that are genetically correlated with MDD don’t have improved power to detect genetic variants in GWAA using the current sample size.

Despite outperforming single traits in endophenotype criteria and ranking methods, composite traits performed more poorly in genome-wide association analysis in terms of the number of more extreme test statistics yielded for each trait, suggesting that improved performance in PRS analysis does not necessarily correspond to increased success in GWAA.

Twenty-two SNPs overlapped with a known gene or its associated regulatory region (19 mapping to protein coding regions, 3 mapping to lincRNA regions). These SNPs were further explored using the GeneCards database to assess the gene(s)’ function and known disease associations. Four genes were mapped to in the binary MDD analysis: WNT11, CACNB2, MAST4, PPFIBP1; two genes for EPQN: CDH23, ENSG00000249209; three genes for GHQ: PWRN1, LRP1B, ECM1; three genes for MDQ: SOD1, DOCK2, SCAF4; four genes for SPQ: MYO3B, C1orf101, GPR137B, ENSG00000244650; two genes for SoS: ECM1, CDH23; and two genes for Gen3: ARAP2, GRM7. No SNPs in Gen4 mapped to a genic or regulatory region.

Interestingly, there was no overlap in gene discovery between binary and quantitative measures of depression suggesting there may be biological utility in our approach. Some of these genes will be analysed in more depth in the discussion section. Summary statistics and biological annotation for SNPs with P-value ≤ 1E-05 are shown in Supplementary Table 5.

### Replication in UKB

To assess whether measures of MDD (binary and quantitative) had a genetic correlation with MDD status in an independent sample, the genetic correlation (r_g_) between GS GWAA summary statistics and MDD GWAA summary statistics from UK Biobank was calculated using bivariate LD Score regression (Bulik-Sullivan, Finucane, et al., 2015b).

Five traits had a high r_g_ (≥+/-0.69) which was within bounds (value between -1 and 1) with UKB MDD. Four traits had a positive correlation: EPQN, GHQ, SoS, and Gen3.

Gen4 had a negative correlation. However, only EPQN and SoS remained statistically significant after multiple testing correction (p≤0.006).

The genetic correlation for MDD in GS and UKB was >1 and out of bounds (r_g_ = 1.96). This can occur if one or both of the traits have a near-zero or negative heritability estimate, which is the case for MDD in GS (h^2^ = 0.04; SE = 0.07). Standard errors are large for all correlations, this could be reduced by using imputed data to increase the number of SNPs used in the analysis.

Results from bivariate LD Score regression are shown in Table 5.

**Table 5.** Bivariate LD Score regression between MDD and candidate endophenotypes in Generation Scotland and MDD in UK Biobank. P-values which survived multiple testing correction are highlighted in bold. r_g_ (SE) denotes the estimated genetic correlation between traits its corresponding standard error. P-value denotes association P-value for the rG estimate; h2_liab (SE) denotes the liability heritability estimate and its corresponding standard error; h2int denotes the single-trait LDSR incept and its corresponding standard error; gcov_int denotes the cross-trait LDSR intercept.

| Trait | $r_g$ (SE) | P-value | $h^2_{liab}$ (SE) | $h^2_{int}$ (SE) | $gcov_{int}$ (SE) |
| --- | --- | --- | --- | --- | --- |
| MDD | 1.91 (1.67) | 0.25 | 0.04 (0.07) | 0.994 (0.007) | -0.007 (0.005) |
| **EPQN** | **0.73 (0.23)** | **0.002** | **0.09 (0.03)** | **1.00 (0.008)** | **-0.005 (0.005)** |
| GHQ | 0.76 (0.38) | 0.04 | 0.04 (0.03) | 1.01 (0.008) | -0.003 (0.005) |
| MDQ | 0.41 (0.57) | 0.47 | 0.03 (0.05) | 1.00 (0.008) | 0.005 (0.006) |
| SPQ | 0.04 (0.27) | 0.895 | 0.09 (0.05) | 0.99 (0.007) | 0.006 (0.005) |
| **SoS** | **0.83 (0.29)** | **0.005** | **0.07 (0.03)** | **1.008 (0.008)** | **-0.005 (0.005)** |
| Gen3 | 0.88 (0.60) | 0.14 | 0.05 (0.06) | 1.004 (0.008) | -0.002 (0.005) |
| Gen4 | -0.69 (0.51) | 0.18 | 0.05 (0.06) | 1.002 (0.008) | -1.99e-04 (0.005) |

## Discussion

In the current study, we sought to aid genetic discovery for depression by revisiting the binary phenotype and developing a quantitative trait using data from Generation Scotland. Our analyses aimed to test whether this derived quantitative trait has improved statistical power to identify genetic risk variants for depression, relative to the binary classification of case/control.

## Summary of findings

### Did quantitative traits fulfil endophenotype criteria?

Two traits, GHQ and EPQN, fulfilled all endophenotype criteria: heritable, genetically and phenotypically correlated with depression, state independent, co-segregating with illness in families, and observed at a higher rate in unaffected relatives than in unrelated controls. A further three traits, MDQ, SPQ, and DSC fulfilled all endophenotype criteria except sharing a statistically significant phenotypic correlation with MDD. Although DSC was a considered as a potential candidate, there was a notable drop in strength of association between DSC and MDD relative to other high-ranking traits (supplementary tables 1, 3, and 4), hence it was not taken forward to create composite measures, which incorporated two (SoS), three (Gen3), or four traits (Gen4) traits. All composite traits fulfilled endophenotype criteria.

### Did endophenotypes “outperform” binary MDD?

Endophenotypes were ranked using three methods (coheritability, variance explained by MDD PRS, and biological insights from GWAA) to assess their statistical power relative to binary MDD.

### Coheritability

Coheritability assesses the covariation between two traits and evaluates the ability of quantitative traits to predict disease liability. Endophenotypes are considered a better predictor of disease liability than the observed trait itself if the trait has a higher coheritability value than the heritability of the disease trait. We considered coheritability using MDD heritability estimates from both the liability (Fernandez-Pujals et al., 2015) and observed scale, as disease genotype is being predicted from case control status and the observed heritability takes account of the loss of information. Gen3 and Gen4 had higher coheritability estimates on the observed scale than MDD, suggesting that these composite traits may be better predictors of disease liability. No trait’s coheritability estimate exceeded the value for MDD on the liability scale.

### MDD PRS

Variance explained by MDD polygenic risk score was chosen as a method to rank candidate endophenotypes because this method utilizes only common trait-associated variants. Therefore it was hypothesized that selecting quantitative traits for use in GWAA based on this method may prove advantageous for gene-discovery.

The two univariate endophenotypes, GHQ and EPQN, and two composite endophenotypes, SoS and Gen3, which survived multiple testing correction all had 2- 3-fold higher trait variance explained relative to binary MDD. One trait which was not taken forward as an endophenotype, LM2D, also survived multiple testing correction, although the variance explained for this trait (0.09%) was very similar to binary MDD (0.08%).

### GWAA

The ultimate purpose of ranking endophenotypes and generating composite measures of depression was to see what we insights we could gain from GWAA, as when these analyses were performed there were only two robustly replicated GWAA hits for depression, identified in the CONVERGE study (Cai et al., 2015).

The majority of top-ranking SNPs did not tag genes which were explicitly linked to psychiatric phenotypes. Four of the 22 genes highlighted in the 8 GWAA we performed had a direct or plausible indirect link to the psychiatric literature: WNT11 and CACNB2 from the MDD analysis, SOD1 from the univariate MDQ analysis, and GRM7 from the composite Gen3 analysis.

Whilst WNT11 itself has not been previously reported in psychiatric disorders, it is inhibited by the Interleukin 1 beta (IL1β) gene which encodes for the IL-1β cytokine protein (Ryu and Chun, 2006). This protein has been previously implicated in psychiatric disorders including association with MDD symptoms in maltreated children of pre-school age (Ridout et al., 2014), association with case-control status in a small schizophrenia case-control study (Kapelski et al., 2015), and increased activity in the bilateral frontal region of the brain, such as the pre-frontal cortex, in schizophrenia patients (Fatjó-Vilas et al., 2012).

CACNB2 encodes for the voltage-dependent L-type calcium channel subunit beta-2 protein and has been cited numerous times in the literature for its potential involvement in the aetiology of psychiatric disorders. SNPs within CACNB2 achieved genome-wide significance in the PGC cross-disorder association meta-analysis of autism, ADHD, bipolar disorder, MDD and schizophrenia (Smoller, 2013) and is associated with bipolar disorder in GWAS in Han Chinese (Lee et al., 2011) and Taiwanese (Jan et al., 2014) populations. Hypermethylation of CACNB2 has been observed in haplotype carriers compared to married-in controls in a Scottish family multiply affected by bipolar disorder and MDD, who carry an illness-linked haplotype on chromosome 4p (Walker et al., 2016).

SOD1 (Superoxide Dismutase 1) encodes an enzyme which catalyzes the dismutation of superoxide anions, a type of reactive oxygen species (ROS). In mice, transgenic overexpression of SOD1 resulted in increased resilience to glucocorticoid-induced depressive-like behaviour relative to wild type mice in the social interaction test, sucrose preference test and forced swim test. Overexpression of SOD1 also resulted in the reduction of a marker of oxidative stress, suggesting that this increased resilience is mediated by decreasing cellular ROS levels (Uchihara et al., 2016). In humans, significantly reduced expression of SOD1 has been observed in conjunction with telomere shortening in white matter oligodendrocytes from MDD donors, relative to control donors (Szebeni et al., 2014). MDD patients have also been shown to exhibit significantly higher activity levels of SOD1 during an acute depressive episode relative to healthy controls (Gałecki et al., 2009).

GRM7 (Metabotropic glutamate receptor 7 precursor) has previously been association with MDD via linkage analysis in a large study of families with severe, recurrent MDD (Breen et al., 2011) and a smaller study of families of heavy smokers. This gene has also been implicated as a potential candidate gene for MDD (p=1.11E- 06) via meta-analysis of three MDD case-control cohorts of European ancestry: STAR*D, GenRED, and GAIN-MDD (Shyn et al., 2011). Interestingly, this region of the genome was also identified for MDD in GS in a GWAS of a continuous measure of MDD created by performing Factor Analysis on SCID questions (unpublished).

### Did findings replicate in UKB?

Only EPQN and SoS survived multiple testing correction in the bivariate LD Score Regression analysis with UK Biobank MDD. Three other traits with a high genetic correlation (GHQ, Gen3, Gen4), which did not survive owing to substantial standard error values. This is due to the GS data not being imputed, as the genetic correlation value is estimated using only SNPs which are in common between GS, UKB, and the LD reference panel (N=460,951). This makes it difficult to definitively ascertain whether the highest ranking endophenotypes in GS correlate with MDD in UKB.

## Limitations

Whilst the question posed in this manuscript is sensible statistically and genetically, the tools and methods used to answer the question were subject to several limitations which are important to consider.

### Pedigree versus genotype methods

Given our outcome of interest was number of SNPs identified via GWAA and their biological interpretation, it may have proven advantageous to use SNP-based metrics of heritability and genetic correlation, instead of pedigree-based methods which may be inflated by capturing non-additive and rare genetic effects.

Indeed, the convergence of our two endophenotype ranking methods - the coheritability and variance explained by MDD PRS were weakly correlated (adjusted R^2^ = 0.09) and non-significant (F_(1,8)_ = 1.9; p = 0.20), as shown in Figure 3.

**Figure 3.**
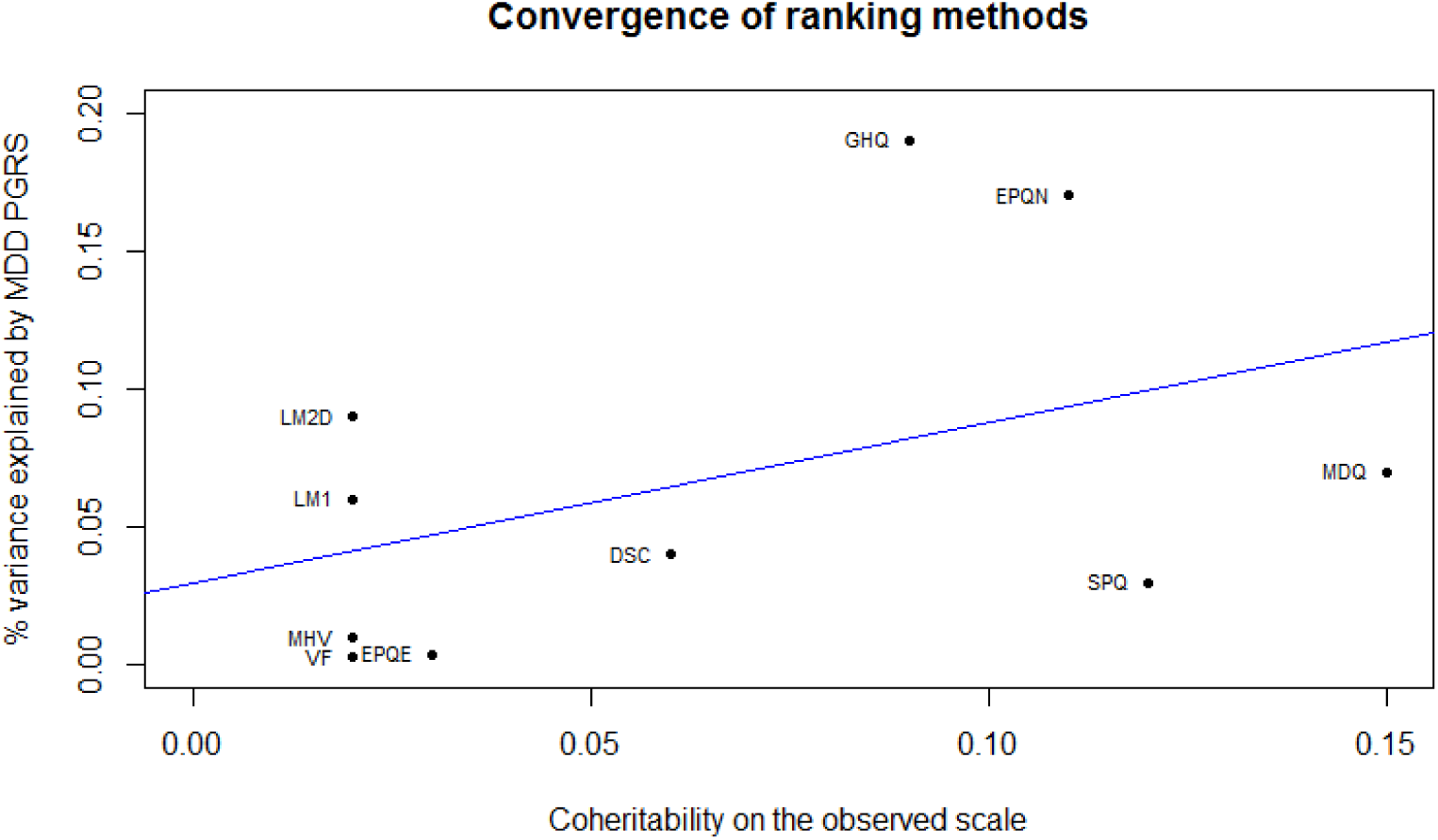
Linear relationship between variance explained by MDD PRS and observed coheritability with MDD for selected mood, personality and cognitive traits, showing a lack of convergence between ranking methods.

However, the choice was made to pursue pedigree-based methods on the basis that heritability estimates using pedigree information do not suffer downward biases as a product of phenotypic heterogeneity to the same extent as estimates using genomic information (Wray and Maier, 2014).

Secondly, at the time this study was conducted, only the raw genotype information was available. Estimates derived from genotype data are limited by the fact that they only account for the variance explained by common SNPs that are tagged by genotyped variants, and not all causal variants, and as such gives a lower bound heritability estimate. This is incongruent with the purpose of this study, namely, to derive a quantitative trait where the heritability was maximized. If the study was repeated in a large, population-based sample with imputed genetic data, using genotypic metrics of coheritability would be the sensible approach.

### Trait selection

EPQN, GHQ, MDQ and SPQ were taken forward as the highest ranking endophenotypes to be used in the creation of composite traits. Although their high genetic correlation and coheritability made MDQ and SPQ an appealing choice, these traits had a markedly reduced sample size (N∼10,000), about half that of other available measures (N∼20,000). It could be fruitful to derive a composite trait which used information from EPQN, GHQ and DSC, as DSC generally performed well across endophenotype criteria, and has the advantage of a sample size more equivalent to EPQN and GHQ.

### Model selection

The mixed family and population structure of GS yields some analytical advantages. For example, complex pedigrees improve the ability to partition variance into causal components, which can be difficult due to the differential rate of decay of environmental and genetic sources of correlation (Tenesa and Haley, 2013). However, this also makes building a regression model which balances adequately accounting for sources of confounding with computational and statistical issues challenging.

In pedigree-based analyses, shared household and sibling environment were fitted as random effects to account for shared environment. The addition of common spouse environment was also tested, and improved model fit for seven traits (GHQ, MDQ, SPQ, DSC, EPQE, LM2D and VF). We chose not to include spouse as a random effect, as without longitudinal data it is difficult to distinguish between common environment and assortative mating. The purpose of fitting environmental sources of resemblance in this study is to ensure that heritability estimates are not overestimated and fitting a spousal environmental effect in the presence of assortative mating would incorrectly partition some of the variance from the genetic effect. However, assortative mating could be explicitly tested using the methods outlined in Robinson et al (Robinson et al., 2017).

#### The way forward

The two primary factors which have hindered genetic discovery in depression have been statistical power and heterogeneity. That being said, since this work was conceived, phenomenal progress has been made in psychiatric genetics. The most recent MDD GWAA employed a minimal phenotyping approach to achieve a substantial discovery sample size (N=807,553) which identified associations in 102 independent loci, 87 of which replicated in an independent sample. This demonstrates that, even without addressing heterogeneity, with considerable sample size increases we gain sufficient statistical power to detect the many variants of small effect (OR = 0.97-1.05) contributing to the polygenic burden of depression and enriched in biological plausible pathways associated with synaptic structure and neurotransmission (Howard et al., 2019).

Focussing on minimal phenotyping has its statistical advantages, but some of the nosological concerns are analogous to the pleiotropic considerations in the current study. Cai and colleagues conducted a study to empirically test the genetic differences between minimal phenotyping and clinically defined depression in the same individuals using UK Biobank data. Their concern with minimal phenotyping is that it introduces substantial measurement error due to the lack of clinical interaction in 50% of self-declared patients, and the prevalence of anti-depressant usage for conditions other than depression. They observed that strictly defined MDD had a higher heritability estimate (h^2^=0.26) than minimal phenotyping (h^2^=0.14), and that the signal for the minimal phenotype was spread across a greater proportion of the genome (80.2%) than strict MDD (65.8%) (Cai et al., 2019).

One could argue that the strong genetic correlation (r_g_=0.86, SE=0.05) between self-reported and clinically diagnosed MDD (Howard et al., 2018), or our quantitative traits and MDD, represents a shared additive component which should ameliorate concerns over phenotypic validity. However, each trait still has a non-shared genetic component. Therefore it does not necessarily follow that signal arising from the analysis of minimal or quantitative phenotypes is representative of what is shared with clinically defined depression. One means of combating this issue would to be to employ methods such as GSEM (Grotzinger et al., 2019) or GWIS (Nieuwboer et al., 2016) to capture the shared genetic component between the trait of interest and clinical depression whilst discarding the non-shared components, then take this factor (GSEM) or principal component (GWIS) forward for analysis.

Regarding heterogeneity, one approach is to stratify the depressed phenotype into discrete subgroups. Recent work by Howard et al attempted this aim using traits which were significantly correlated with depression and sufficiently well powered for analysis using BUHMBOX. They observed no evidence for subgroups within depression with the psychiatric, autoimmune, and anthropomorphic traits tested, suggesting the genetic correlations between these traits and MDD were driven by pleiotropic variants carried by most or all cases, rather than a specific subgroup (Howard et al., 2020).

So the question remains - how can we exploit pleiotropy to characterize the biology of depression? One possibility would be to identify endophenotypes of specific symptom domains, as individual symptoms are differentially heritable (Fried and Nesse, 2015a) and risk factors exert differential influences on individual symptoms (Lux and Kendler, 2010; Fried et al., 2014). This would prevent approaching depression as a sum-score of many disparate symptoms, collapsing individuals into one undifferentiated category which assumes that symptoms are interchangeable and equal in pathological contribution, when that is not the case (Fried and Nesse, 2015a). For the purposes of case control studies, it would also be invaluable for control participants to fully complete structured interviews, as a lot of valuable information is lost due to questions being skipped once the initial screening questions have been administered. This additional information could be used to inform stratification, with multivariate methods such as item response theory and structural equation modelling being employed to analyse the exact relationships between symptoms and underlying dimensions (Fried and Nesse, 2015a).

## Conclusions

When considering the evolution of our understanding of inheritance, the plant hybridization experiments of Gregor Mendel are forefront in our consciousnesses. Mendel’s observations were undoubtedly extraordinary and revolutionary, establishing that two parental units of inheritance give rise to phenotypic traits - nearly a century before the characterization of DNA. However, the binary lens through which traits were viewed gives rise to a false model of dominance, encouraging a reductive genetic determinism that has previously been ascribed the moniker “Scientific Calvinism” (Radick, 2016).

Weldon’s revisitation of Mendel’s experiments, albeit much less remembered, demonstrate theories that are a much closer approximation to how we understand complex trait genetics today - embracing complexity and variation. If we continue to categorize quantitative liabilities as binary traits, we risk deliberately excluding variability that would produce alternative pathophysiological associations (Stearns, 2010), and being limited to only observing what has already been observed.

Mendel’s ratio and the co-segregation of traits would remain consistent through the generations if the traits remained binary, yet no single gene has been identified as having a pleiotropic effect on seed coat colour, flower petal colour, and axial spots. So whilst Mendelian genetics plays a historical, foundational role in how we approach the biggest challenges of psychiatric genetics today, the Weldonian emphasis on variation, ancestry, and environment is much more likely to help us bridge the gap between statistical association and functional consequences.

## Data Availability

Data are available to qualified researchers on a cost-recovery basis via online application processes, accessible via www.gsaccess.org and www.ukbiobank.ac.uk/register-apply/

## Acknowledgements

Generation Scotland received core funding from the Chief Scientist Office of the Scottish Government Health Directorate CZD/16/6 and the Scottish Funding Council HR03006. Genotyping of the Generation Scotland samples was carried out by the Genetics Core Laboratory at the Wellcome Trust Clinical Research Facility, Edinburgh, Scotland and was funded by the UK’s Medical Research Council and the Wellcome Trust (Wellcome Trust Strategic Award “STratifying Resilience and Depression Longitudinally” (STRADL) (Reference 104036/Z/14/Z). YZ acknowledges support from China Scholarship Council. AMMcI acknowledges support from the Dr Mortimer and Theresa Sackler Foundation. This work has made use of the resources provided by the Edinburgh Compute and Data Facility (ECDF). (http://www.ecdf.ed.ac.uk/).

We are grateful to all of the individuals who participated in Generation Scotland and UK Biobank, the general practitioners and research staff for their help in recruiting them, and the whole Generation Scotland team, which includes interviewers, computer and laboratory technicians, clerical workers, research scientists, volunteers, managers, receptionists, healthcare assistants and nurses.

Lynsey S. Hall would like to extend additional thanks to former colleague and office mate Dr Toni-Kim Clarke, whose relentless encouragement and friendly berating ensured the completion of the PhD project analyses presented in this paper; and to Dr Antonio F. Pardiñas, for his insightful reading list suggestions from the biometrician-Mendelian era.

## Author contribution statement

Lynsey S. Hall performed all analyses in the current manuscript unless otherwise specified, interpreted the results, and wrote the research manuscript.

Mark J. Adams ran performed confirmatory quality control checks on the phenotype and genotype data from UKB, and ran the genome-wide association analysis of MDD in UKB.

Yanni Zeng created the environmental matrices used in pedigree-based analyses.

Jude Gibson and Ella M. Wigmore generated the polygenic risk scores in GS and UKB.

Ana Maria Fernandez-Pujals generated the liability-scale MDD heritability estimate and was involved with quality control of the GS phenotype data.

Generation Scotland for generating the data performing initial quality control of the genotype data.

Heather C. Whalley, Chris S. Haley, and Andrew M. McIntosh supervised the work. They were instrumental in the study design, in establishing appropriate statistical modelling and in interpreting the results.

## Figure legends

Tables and/or Figures (uploaded separately)

